# Genetic and lifestyle modifiers of haemochromatosis-related clinical outcomes in HFE C282Y homozygotes: prospective cohort study in UK Biobank

**DOI:** 10.1101/2025.08.22.25334187

**Authors:** Mitchell R Lucas, João Delgado, Robin N Beaumont, Gareth Hawkes, Andrew R Wood, Caroline F Wright, Jeremy Shearman, Janice L Atkins, Luke C Pilling

## Abstract

**Background:** The iron overload disease haemochromatosis is primarily caused by *HFE* p.C282Y homozygosity, yet penetrance of clinical outcomes (including liver disease/cancer) varies. We estimated the effect of genetic and lifestyle factors on disease penetrance and expressivity in C282Y homozygotes.

**Methods:** We analysed 2,893 C282Y homozygous UK Biobank participants (n=1,295 male). We ascertained haemochromatosis from medical records, liver disease/cancer, osteoarthritis, joint replacement surgeries, and dementia diagnoses. We derived polygenic scores (PGS) for iron biomarkers including hepcidin and transferrin saturation (TSAT). Sex-stratified logistic regression assessed associations with clinical outcomes. We used time-to-event regression estimating effects of age, lifestyle, and PGS. We estimated effects of rare *HFE* variants using whole genome sequencing data.

**Results:** In male *HFE* C282Y homozygotes, higher TSAT PGS increased likelihood of diagnosis of haemochromatosis, and separately any clinical consequence (OR_top-vs-bottom-PGS-quintile_=1.83, 95%CI: 1.26-2.66, p=0.001). Cumulative incidence of assessed haemochromatosis clinical outcomes in men by age 80 was 64.5% (highest quintile) versus 51.6% (lowest) (p-value for difference=0.025). In females, TSAT PGS increased haemochromatosis likelihood (cumulative incidence: 45.3% vs. 23.3% [highest/lower quintile], p=0.00001) but not liver disease. PGS for other iron biomarkers were not significantly associated with clinical outcomes. Rare heterozygous predicted loss-of-function variants in *HFE* increased haemochromatosis likelihood in non-C282Y homozygotes (aggregate-OR=14.8, 95%CI 4.7-41.1, p=0.003), highlighting the importance of sequencing undiagnosed individuals to find rare causes of haemochromatosis.

**Conclusion:** Higher genetically predicted transferrin saturation significantly increased risk of clinical outcomes in *HFE* C282Y homozygotes. Combined with modifiable lifestyle factors, genetic information could refine risk stratification and personalise iron monitoring, following validation.

## 1 Introduction

HFE haemochromatosis is the most common genetic disorder among individuals of Northern European ancestry.(1) It is predominantly caused by *HFE* C282Y homozygosity, seen in 1 in 150 individuals of estimated European (EUR) genetic ancestry (C282Y allele frequency = 7%).(2) While the *HFE* H63D variant is more common (allele frequency 10-29%), its clinical manifestations are much lower.(3) Haemochromatosis is characterised by iron overload which significantly increases the risk of clinical outcomes,(4) including liver disease and hepatocellular carcinoma,(5) musculoskeletal disorders,(6) neurodegenerative diseases,(7) and all-cause mortality.(8) However, clinical penetrance among C282Y homozygotes varies.(9,10) In UK Biobank participants of EUR-like genetic ancestry, we estimated that 56% of C282Y homozygous males and 41% of C282Y homozygous females are expected to be diagnosed with haemochromatosis by age 80.(8) Though this is higher than many previous estimates, it suggests other factors (genetic or lifestyle) modify phenotypic expression within C282Y homozygotes.(11,12)

HFE C282Y results in reduced hepcidin and increased iron absorption. Other iron metabolism genes include HAMP, BMP2, FTL, SLC40A1, TMPRSS6, and TF. Genetic variants in these genes are known to increase serum iron status biomarkers such as serum ferritin (SF), transferrin saturation (TSAT), and iron.(13–18) Notably, TF polymorphisms explain around 40% of transferrin variation and mutations in HJV impair BMP signalling,(18) reducing hepcidin levels and can cause severe iron overload.(19) We previously reported increased risk of liver disease in UK Biobank participants with higher genetically predicted TSAT.(11) Additionally, we demonstrated that lifestyle factors and patient characteristics, such as alcohol intake, smoking, frequent consumption of red or processed meat, and obesity, were associated with higher levels of MRI-estimated liver iron in ∼40,000 UK Biobank participants.(12)

In this study, we created weighted allele scores (polygenic scores, [PGS]) for each participant to estimate their genetically predicted iron metabolism biomarkers, including novel predictors hepcidin, serum transferrin receptor (sTfR), as well as serum iron, TSAT, SF, and total iron-binding capacity (TIBC). This study substantially extends previous work by incorporating a longer follow-up and expanding the risk factors, genetics, and clinical outcomes examined. Using data from the UK Biobank, we aimed to clarify genetic and lifestyle modifiers of clinical penetrance in *HFE* C282Y homozygotes. By using PGS related to iron metabolism biomarkers and key lifestyle characteristics, we quantified their combined predictive value for clinically meaningful outcomes. We also investigated the effect of rare genetic mutations in *HFE* in non-C282Y homozygotes, aiming to determine novel genetic risk factors for haemochromatosis.

## 2 Methods

### 2.1 Study population

UK Biobank includes 502,464 community volunteers with baseline ages 39 to 73 years, recruited from 22 centers in the UK (England, Scotland, and Wales) from 2006 to 2010. Participants were moderately healthier than the general population(20); however, *HFE* allele frequencies were comparable to other UK-based studies.(5) Our studied sample included participants (n = 450,401) genetically similar to the 1000 Genomes Project European Ancestry superpopulation (‘EUR-like’),(21) the definition of this population has been described previously.(22) *HFE* C282Y (rs1800562 G>A; NM_000410.3(HFE):c.845G>A [p.Cys282Tyr]) and H63D (rs1799945 C>G; NM_000410.4(HFE):c.187C>G [p.His63Asp]) genotype information were from the whole genome sequencing DRAGEN variant calls (UK Biobank field: 24310 (23)). UK Biobank obtained ethics approval from the Northwest Multicenter Research Ethics Committee (reference: 11/NW/0382). Patient consent did not include patient feedback to participants on genotype. Access was granted under UK Biobank approved applications 14631 and 103356.

### 2.2 Baseline assessment (2006–2010)

We calculated body mass index (BMI) as weight (kg, field: 21002) divided by height (m^2^, field: 50), waist-hip-ratio (WHR) by dividing waist circumference (cm, field: 48) by hip circumference (cm, field: 49). WHR was categorized into a binary variable based on sex-specific thresholds for central obesity.(24) Participants with WHR ≥0.96 (males) or WHR ≥0.85 (females) were assigned a value of 1, indicating high WHR, while those below these thresholds were assigned 0. Alcohol intake (fields: 1568 [weekly red wine], 1578 [weekly white wine/champagne], 1588 [weekly beer/cider], 1598 [weekly spirits], 1608 [weekly fortified wine], 4407 [monthly red wine], 4418 [monthly white wine/champagne], 4429 [monthly beer/cider], 4440 [monthly spirits], 4451 [monthly fortified wine], 4462 [monthly other alcoholic drinks], 5364 [weekly other alcoholic drinks])) was categorised by the number of units drank per week groups into ‘0 units per week’, 1-14 units per week’ (reference group, based on UK national recommendations1), ’15-29 units per week’ and ‘over 30 units per week’. Smoking status (field: 20116) was defined as ‘current smoker’ based on subjects being asked “Do you smoke tobacco now? And grouped in a binary variable of 0 = no and 1 = yes, on most or all days’ and ‘Only occasionally’. Each participant was asked separate questions about their meat consumption during the assessment. They were specifically asked how often they ate processed meat (field: 1349), lamb/mutton (field: 1379), pork (field: 1389), and beef (field: 1369). For each type of meat, participants could choose from the following options: ‘Never,’ ‘Less than once a week,’ ‘Once a week,’ ‘2-4 times a week,’ ‘5-6 times a week,’ or ‘Once or more daily’. We provided a value for each respective response on meat consumption: (Never = 0) (Less than once a week = 0.5) (Once a week = 1) ( 2-4 times a week = 3) (>4 times a week = 5.5) then derived a summed total weekly consumption of red /processed meat and recorded as follow ‘0 times/week’ (reference group) and ‘0.1-2.9 times/week’, and ‘≥3.0 times/week’.(25)

### 2.3 Diagnosis ascertainment

England, Wales, and Scotland hospital inpatient admission records (Hospital Episode Statistics [HES]) were available from April 1996 to October 2022. Incident diagnoses and surgical procedures were from hospital inpatient data (baseline to October 2022) plus cancer registries to December 2020 for England and Wales and November 2021 for Scotland. Corresponding primary care diagnosis codes were identified using the UKB “Clinical Coding Classification Systems and Maps” resource to map ICD-10 codes to Read2/CTV3 (UK Biobank resource: 592). Outcomes ascertained were haemochromatosis, liver disease, liver fibrosis or cirrhosis, liver cancer, osteoarthritis, joint replacement surgeries, dementia, and Alzheimer’s Disease, non-Alzheimer’s dementia (see supporting information [SI] Table 1 for ascertainment codes for all outcome diagnoses). Prevalent diagnoses also included data from baseline self-report information (fields: 20001, 20002). We used R package {ukbrapR} v0.3.5 to ascertain diagnoses in the UK Biobank Research Analysis Platform (https://github.com/lcpilling/ukbrapR). “Joint replacement surgeries” included hip, knee, ankle, or shoulder replacement surgery.

**Table 1:**
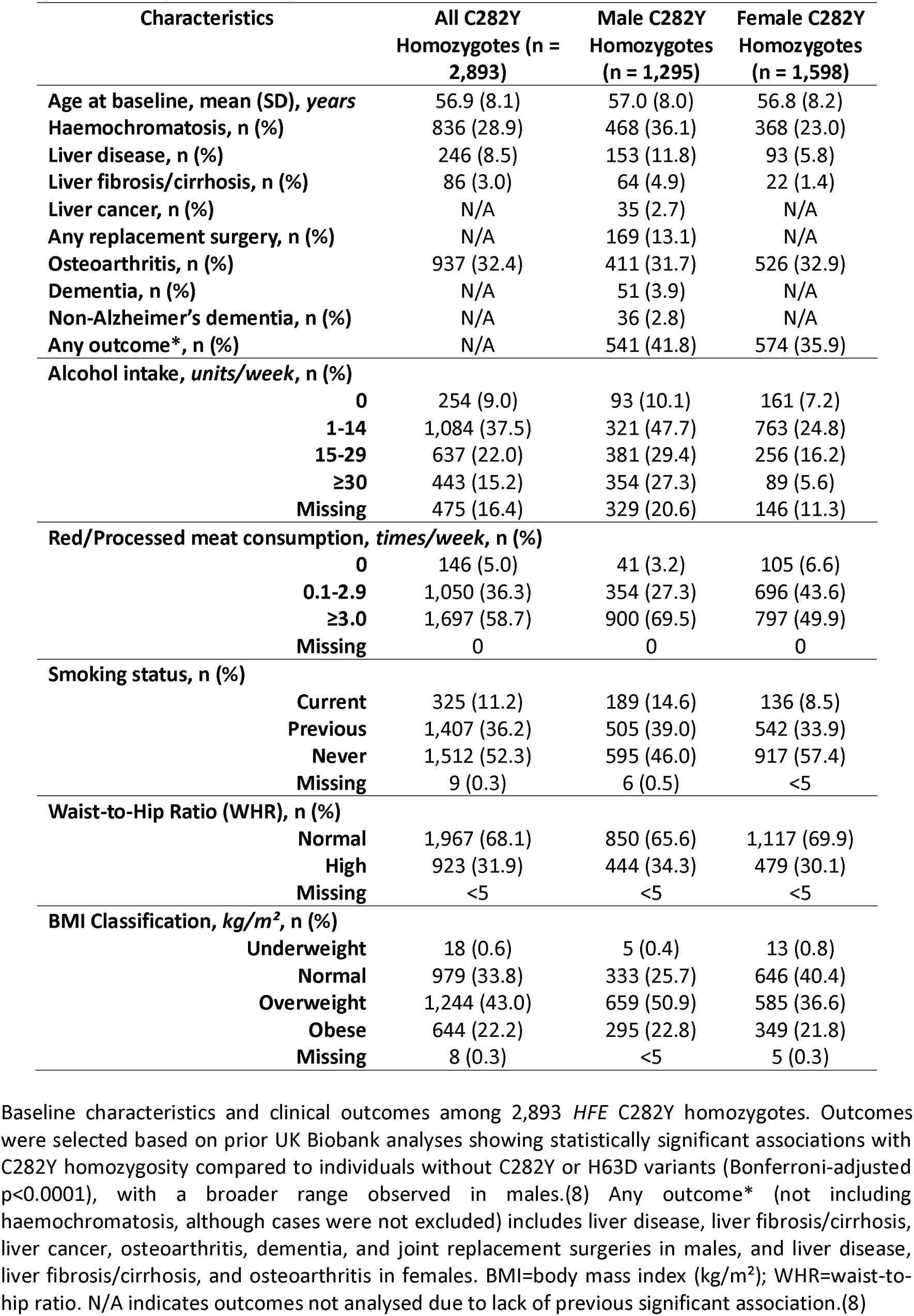
Baseline characteristics of the UK Biobank *HFE* C282Y homozygote participants of European ancestry, stratified by sex.

### 2.4 Polygenic Score for Iron Biomarkers

We used data from the UK Biobank whole genome sequencing DRAGEN variant calls (BGEN format, field: 24309)(23) to create PGS for six iron-status biomarkers including hepcidin, sTfR, serum iron, TSAT, SF, and TIBC, among 451,046 participants of EUR genetic ancestry. For hepcidin and sTfR, we used 43 independent genetic variants identified in a genome-wide association meta-analysis of up to 91,675 participants for hepcidin and 45,330 participants for sTfR, from 12 European-based cohorts (CHRIS, CROATIA_Vis, Danish Blood Donor Study, deCODE, FinDonor 10,000 Studies 1 and 2, InCHIANTI, INGI-Val Borbera, INTERVAL, KORA_F3, Nijmegen Biomedical Study, and PREVEND). The analysis identified 15 novel loci associated with hepcidin and sTfR concentrations.(26) For serum iron, TSAT, SF, and TIBC we used 128 independent genetic variants identified in a genome-wide association study (GWAS) of 257,953 individuals from six cohorts:(27) the Trøndelag Health Study (HUNT), Michigan Genomics Initiative (MGI), SardiNIA, deCODE (Iceland), Interval Study (UK), and the Danish Blood Donor Study (DBDS). The study population was predominantly of European genetic ancestry. These included 20 variants linked to iron levels, 64 to ferritin, 19 to TSAT, and 41 to TIBC. Given that genetic variants remain stable throughout life, PGS serve as measures of lifelong genetic risk; individuals with higher PGS (e.g., genetically predicted higher serum iron levels) have higher lifetime susceptibility to higher levels.(28) See SI Table 2 for variants included. We used R package {ukbrapR} v0.3.5 to create the polygenic scores in the UK Biobank Research Analysis Platform (https://github.com/lcpilling/ukbrapR). In brief, this uses ‘bgenix’ (https://enkre.net/cgi-bin/code/bgen/doc/trunk/doc/wiki/bgenix.md) to extract the variant calls from the UK Biobank whole genome sequencing DRAGEN variant calls (BGEN format, field:24309), and ‘plink2’ (https://www.cog-genomics.org/plink/2.0/) to convert the subsetted BGEN file to PGEN format and create the weighted allele score.

**Table 2:**
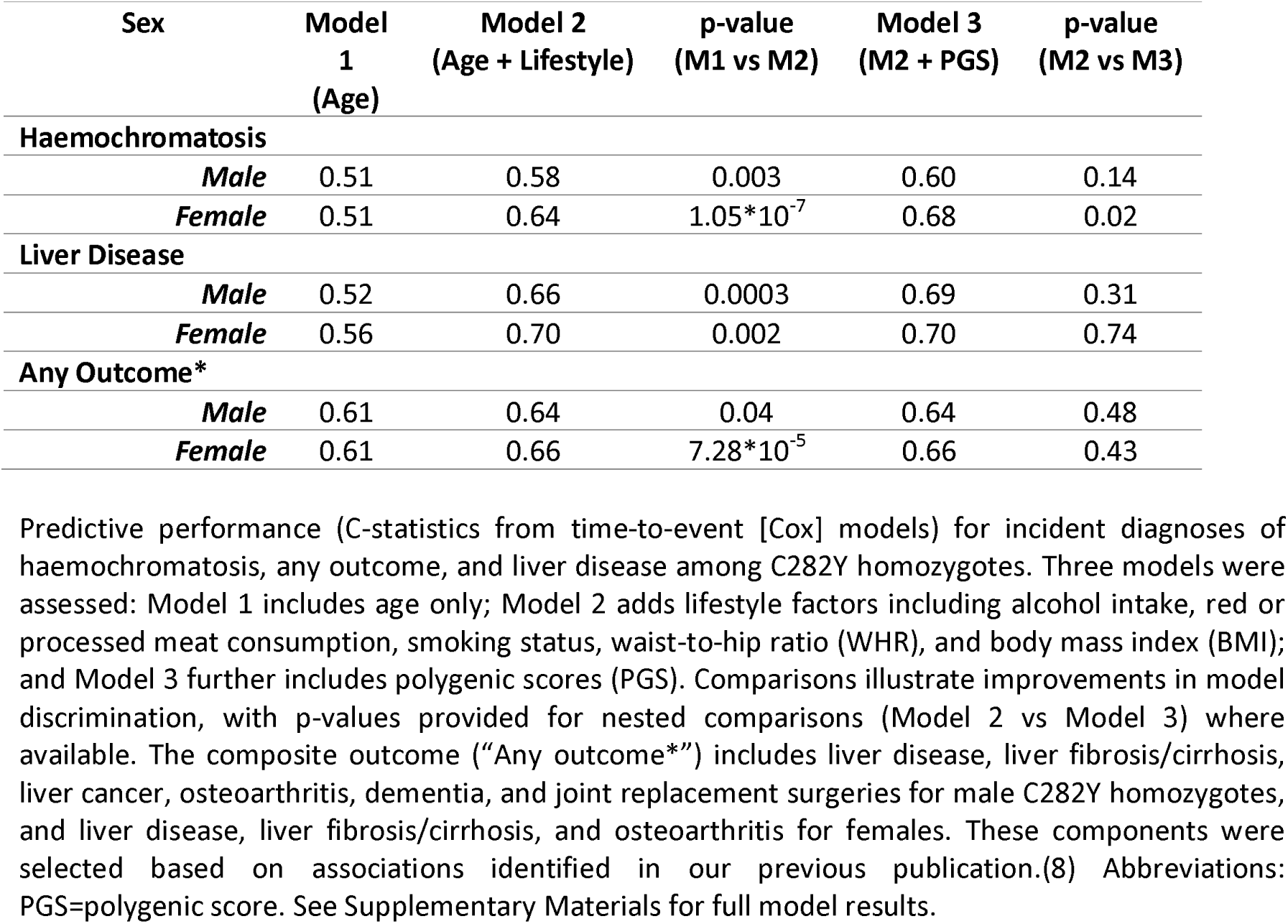
Predictive Model Comparison by Outcome and Sex in *HFE* C282Y Homozygotes.

### 2.5 Analysis of rare variants in HFE

We used data from the UK Biobank whole genome sequencing DRAGEN variant calls (BGEN format, field : 24309)(23) in the 451,046 participants of EUR genetic ancestry and REGENIE v3.3 to estimate associations between genetic variants in the *HFE* gene region (± 2Mbp of the *HFE* gene: human genome build GRCh38 coordinates chr6:24087525-28098445) and diagnosed haemochromatosis, adjusted for age, sex, relatedness, and population stratification. The analysis was performed 1) at the single-variant level for variants with minor allele count ≥10, and 2) by aggregating rare variants (minor allele frequency <0.001) with similar predicted consequence e.g., “predicted loss-of-function” (pLoF’, including frameshift, stop gain, and splice acceptor/donor variants) or “missense” variants (aggregate variant analysis increases power compared to single variant analysis). Sensitivity analyses were conditioned on *HFE* C282Y genotype to adjust for any linkage disequilibrium, i.e., to identify novel haemochromatosis causes beyond the known genotypes.

### 2.6 Statistical Analysis

We first analysed each iron biomarker PGS as a continuous variable within *HFE* C282Y homozygous participants. Each PGS was standardised to z-scores (mean = 0, standard deviation = 1) prior to analyses to ensure interpretability of effect estimates across models and between PGS. Sex-stratified logistic regression models, adjusted for age, assessment center, and genetic principal components (PC1-PC10), were used to evaluate associations between standardized PGS and clinical outcomes. Genetic principal components were included to control for population stratification bias.(29)

Next, *HFE* C282Y homozygous participants were categorised into quintiles based on each iron biomarker PGS. We conducted sex-stratified logistic regression analyses comparing clinical outcomes between the highest and lowest quintiles, adjusting again for age, assessment center, and PC1- PC10.(29)

Clinical outcomes included in this analysis were selected based on our previously published study,(8) which identified specific outcomes that were significantly (FDR-adjusted p < 0.05) increased in risk within male or female C282Y homozygotes compared to individuals without *HFE* C282Y or H63D variants. These were: haemochromatosis, liver disease, liver fibrosis/cirrhosis, liver cancer, osteoarthritis (both sexes), joint replacement surgeries, dementia, and non-Alzheimer’s dementia in male C282Y homozygotes; and haemochromatosis, liver disease, liver fibrosis/cirrhosis and osteoarthritis in females. We aggregated haemochromatosis-associated clinical outcomes into a composite outcome termed “Any outcome” throughout (to clarify, haemochromatosis cases were not excluded from the primary analysis, but the “Any outcome” includes haemochromatosis-associated clinical complications).

In this analysis, we applied Benjamini-Hochberg (BH) False Discovery Rate (FDR) correction. The BH-FDR correction was applied separately to tests conducted in males (n = 54) and females (n = 30), as no direct comparisons between sexes were performed. P-values were ranked, and FDR-adjusted significance thresholds (q = 0.05) were determined to control the expected proportion of false discoveries.

Kaplan-Meier analyzes estimated cumulative incident probabilities for outcomes, comparing the lowest and highest PGS quintiles from age 40-80 years, stratified by sex. To assess differences in cumulative incidence at specific time points (e.g., age 80), we compared survival probabilities between groups using a Z-test based on Greenwood’s standard errors.

For predictive modeling we used time-to-event (Cox regression) models from baseline assessment to the event date or date of censoring (30 October 2022). Participants with prevalent diagnoses of clinical outcomes (assessed previously in logistic regression models) at baseline were excluded to maintain temporality between baseline measurements (lifestyle factors and genetic predictors) and subsequent clinical outcomes, in accordance with Bradford Hill’s temporality criterion for causality.(30) We compared the Harrell’s C-statistic between models to evaluate relative predictive performance of age, iron-related lifestyle factors (alcohol intake, smoking, red/processed meat consumption, WHR, and BMI), and iron biomarker PGS individually and in combination, stratified by sex.

We also performed sex-stratified analysis of serum iron and TSAT polygenic scores on haemochromatosis separately in other *HFE* C282Y and H63D genotype groups (H63D heterozygotes, H63D homozygotes, C282Y-H63D compound heterozygotes, C282Y heterozygotes, and individuals without C282Y or H63D variants).

Unless otherwise specified we used R version 4.1.1 and REGENIE version 3.3 on the UK Biobank Research Analysis Platform for analysis. Primary analyzes were performed between November 2024 to June 2025.

### 2.7 Sensitivity Analysis

In sensitivity analyses we excluded those diagnosed with haemochromatosis at baseline assessment to avoid bias from those likely undergoing iron monitoring and treatment or who may have received treatment shortly after baseline. We also assessed the potential impact of familial relatedness by removing participants related to the third-degree or closer. Finally, associations with rare *HFE* pLoF variants were explored, highlighting their independent contribution to risk of haemochromatosis, liver disease, liver cancer, osteoarthritis, and dementia, adjusting for age, sex, PC1-PC5, and known *HFE* haemochromatosis-associated genotypes (C282Y and H63D).

## 3 Results

### 3.1 Participant characteristics

At baseline, the mean age of the 1,295 male C282Y homozygotes was 57.0 years (SD 8.0). By the medical records censoring date (30 October 2022), 36.1% of males had been diagnosed with haemochromatosis. Based on outcomes previously shown to be significantly associated with C282Y homozygosity in UK Biobank analyzes (Bonferroni-adjusted p1<10.0001), several liver-related conditions were prevalent among males, including liver disease (11.8%), liver fibrosis or cirrhosis (4.9%), and liver cancer (2.7%). Osteoarthritis was reported in 31.7% of males. These outcomes, collectively considered as part of “any outcome” in males, also include dementia and joint replacement surgeries, though their individual prevalence was not reported here. Additionally, males exhibited relatively high alcohol consumption, with 27.3% consuming ≥30 units per week, and higher red or processed meat intake, with 27.3% consuming these foods between 0.1 and 2.9 times weekly. Approximately 46.0% of males had never smoked. At baseline, 50.9% of males were classified as overweight (BMI 25–29.9 kg/m²), while 25.7% maintained a normal BMI (18.5–24.9 kg/m²) (Table 1).

At baseline, the mean age of the 1,598 female C282Y homozygotes was 56.8 years (SD 8.2). By the censoring date (30 October 2022), 23.0% of females had received a haemochromatosis diagnosis. Liver-related conditions among females included liver disease (5.8%), liver fibrosis or cirrhosis (1.4%), and liver cancer (0.3%). Osteoarthritis was reported in 32.9% of females. Alcohol intake among females was 5.6% consuming ≥30 units per week. Females also had lower red or processed meat consumption, with 43.6% reporting intake of these foods between 0.1 and 2.9 times weekly. A majority (57.4%) of females had never smoked. At baseline, 36.6% of females were overweight, and 40.4% had a normal BMI (Table 1).

We assessed multicollinearity among the iron PGS biomarkers using correlation matrices and variance inflation factors (VIFs). Serum iron and transferrin saturation PGSs were highly correlated (r = 0.84), with VIFs of 7.66 and 12.66, respectively (Fig 1). To address this, we excluded serum iron from subsequent multivariable models, based on evidence of high collinearity with transferrin saturation, as indicated by both the correlation matrix and VIF diagnostics (Fig 1). These analyzes were conducted in the full UK Biobank EUR-like sample (n = 451,046).

**Fig 1:**
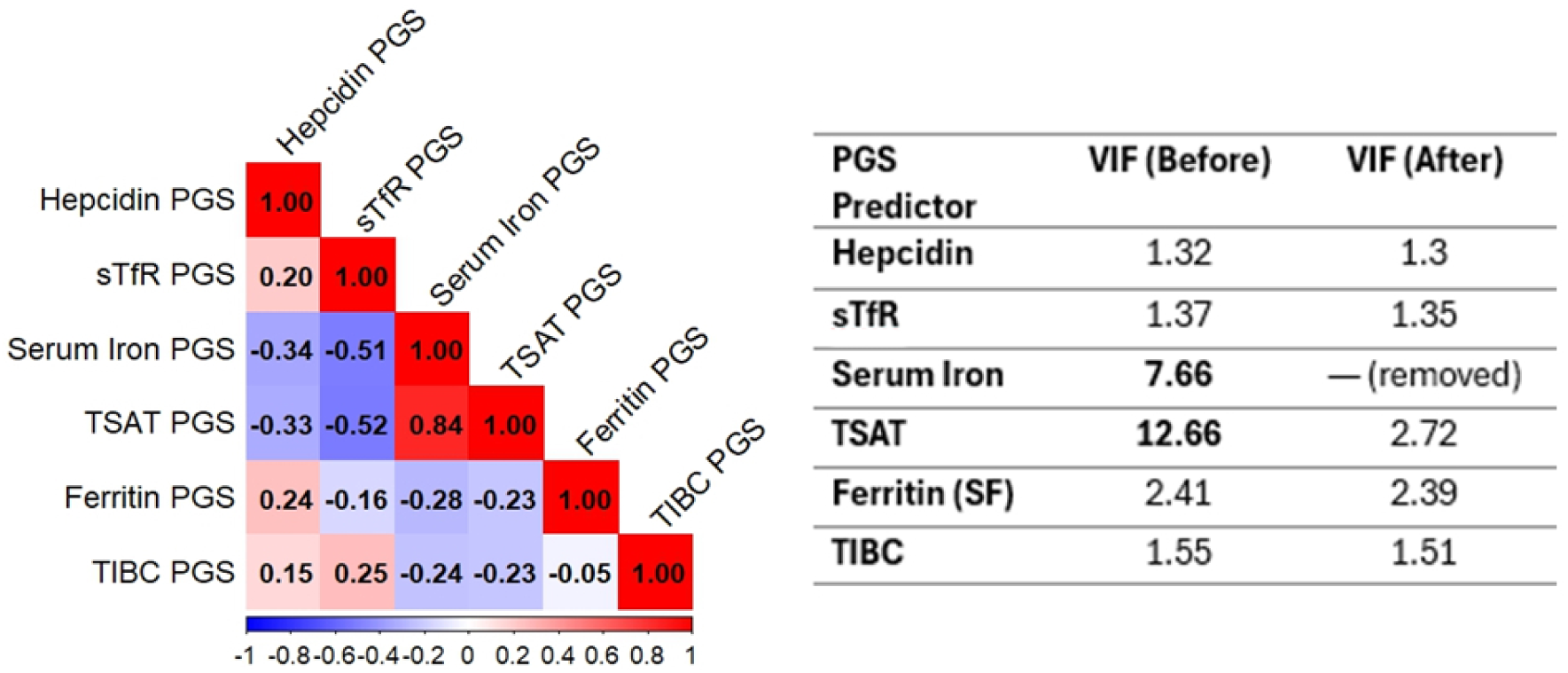
Pearson’s Correlation Between Iron Status Biomarker Polygenic Scores. A) Pearson correlations between polygenic scores for six iron-related biomarkers within the whole UK Biobank EUR-like sample (n=451,046). B) VIF table illustrating multicollinearity diagnostics before and after the removal of serum iron. The decision to exclude serum iron was based on evidence of high collinearity with transferrin saturation, as indicated by both the correlation matrix and VIF values.

After applying the BH FDR correction to account for multiple testing, significance thresholds for analyzes using PGS as a continuous variable for males and females C282Y homozygotes became p ≤ 0.009 and p ≤ 4×10^-6^, based on 54 and 30 independent tests, respectively. See SI Tables 3 and 4 for BH-FDR calculations. Analyzes using PGS as highest vs lowest quintiles were p ≤ 0.0014 for males and p ≤ 6×10^-5^ for females (SI Tables 5 and 6).

### 3.2 PGS and Disease Outcomes in Male *HFE* C282Y Homozygotes

Males with higher TSAT PGS showed significantly increased odds for haemochromatosis diagnosis (OR per 1-SD increase = 1.22, 95%CI: 1.09-1.38 , p=0.0009), liver disease (OR=1.30, 95%CI: 1.09-1.55, p=0.004), liver fibrosis or cirrhosis (OR=1.66, 95%CI: 1.27-2.20 , p=0.0003), and joint replacement surgeries (OR=1.27, 95%CI: 1.07-1.51, p=0.006) compared to those without a diagnosis of these clinical outcomes.

Additionally, increased odds were observed for any haemochromatosis-associated morbidity previously shown to be elevated in *HFE* C282Y homozygous males (liver disease, liver cancer, osteoarthritis, joint replacement surgeries, or dementia)(8) (OR=1.20, 95%CI: 1.07-1.35, p=0.003) (SI Table 7).

Similarly, higher serum iron PGS was strongly associated with increased odds of haemochromatosis (OR per 1-SD increase = 1.30, 95%CI: 1.16-1.47, p=0.00002), liver fibrosis or cirrhosis (OR=1.57, 95%CI: 1.19-2.08, p=0.002), liver cancer (OR=1.75, 95%CI: 1.21-2.57, p=0.004), and joint replacement surgeries (OR=1.32, 95%CI: 1.11-1.57, p=0.002). A significant association was also observed with any haemochromatosis-related outcome (OR=1.22, 95%CI: 1.09-1.38, p=0.0009). Serum iron PGS was not statistically associated with dementia outcomes (SI Table 7).

Hepcidin, soluble transferrin receptor (sTfR), serum ferritin, and TIBC polygenic scores were not associated with outcomes after controlling for multiple tests (p ≤ 0.009).

Males in the highest TSAT PGS quintile (20%) had increased likelihood of haemochromatosis (OR=1.92, 95%CI: 1.32-2.79, p=0.0007) compared to those in the lowest quintile (Fig 2A), with a cumulative incidence by age 80 of 55.4% [95%CI: 46.4%-62.8%] compared to 38.0% [29.4%-45.5%] with a statistical difference of p=0.003 (see SI Table 8 for full list of male cumulative incidence results from age 40 to 80 years). TSAT PGS was also associated with increased likelihood of any haemochromatosis-associated morbidity (a composite outcome including liver disease, liver fibrosis/cirrhosis, liver cancer, osteoarthritis, dementia, non-Alzheimer’s dementia, and joint replacement surgeries)(8) (OR=1.83, 95%CI: 1.26-2.66, p=0.001), with a cumulative incidence of 64.5% [95%CI: 55.9%-71.3%] compared to 51.6% [42.6%-59.3%] in the lowest quintile (p=0.025) (Fig 2A,B and SI Table 8). As shown in both Fig 2A and 2B. In Fig 2A, a forest plot displays the odds ratios for individual hospital-diagnosed clinical outcomes including liver disease, liver fibrosis/cirrhosis, liver cancer, osteoarthritis, dementia, non-Alzheimer’s dementia, and joint replacement surgeries comparing the top versus bottom quintiles of TSAT PGS in male C282Y homozygotes (n = 1,295). Points are color-coded by statistical significance, with red indicating results that survived FDR correction (p ≤ 0.0014), orange for nominal significance (p < 0.05), and grey for non-significant findings (p > 0.05). Fig 2B complements this by showing the cumulative incidence curves for the composite outcome across age 40 to 80 years, demonstrating increasing disease burden over time among those with higher genetically predicted TSAT.

**Fig 2:**
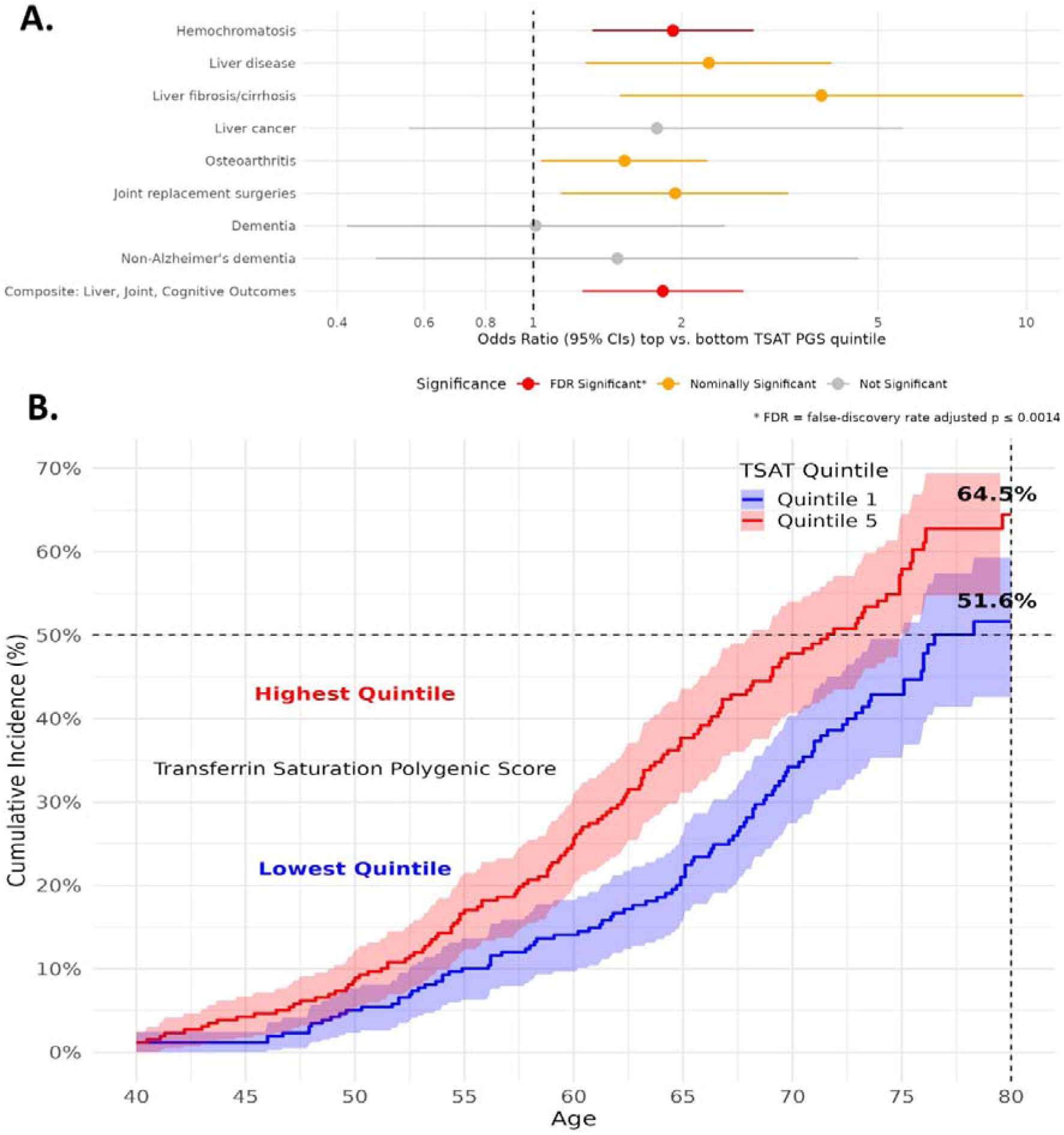
Associations between high genetically predicted transferrin saturation and clinical outcomes in male C282Y homozygotes. A) Forest plot from logistic regression analyses in male C282Y homozygotes (n=1,295) of associations between high genetically predicted TSAT (top PGS quintile) and risk of hospital-diagnosed clinical outcomes (vs bottom PGS quintile). The composite outcome includes liver disease, liver fibrosis/cirrhosis, liver cancer, osteoarthritis, dementia, non-Alzheimer’s dementia, and joint replacement surgeries. Red points indicate associations that passed the Benjamini-Hochberg False Discovery Rate (FDR) threshold (q < 0.05), orange points represent nominal significance (p < 0.05), and grey points are not significant. B) Cumulative incidence for the composite outcome is plotted in the high and low TSAT PGS groups, showing the cumulative incidence of diagnoses from age 40 to 80. See SI Figure 1 for equivalent plots after excluding participants with diagnosed haemochromatosis.

Equivalent cumulative incidence plots excluding participants with a baseline diagnosis of haemochromatosis are provided in SI Fig 1, where effect sizes were slightly attenuated (SI Table 9). Nominal associations (p < 0.05) were observed with diseases individually: liver disease (OR=2.27, 95%CI: 1.28-4.01, p=0.005), liver fibrosis/cirrhosis (OR=3.84, 95%CI: 1.50-9.83, p=0.005), osteoarthritis (OR=1.52, 95%CI: 1.04-2.25, p=0.03), and joint replacement surgeries (OR=1.94, 95%CI: 1.14-3.28, p=0.01) (SI Fig 2); though none remained statistically significant after correction (p ≤ 0.0014).

Similarly, males in the highest serum iron PGS quintile had significantly higher odds of being diagnosed with haemochromatosis (OR=2.13, 95%CI: 1.46-3.10, p=0.00008), with an estimated cumulative incidence to age 80 years of 59.3% [95%CI: 50.6%-66.5%] compared to 37.6% [29.2%- 45.1%] in the lowest quintile (p=0.00015) (See SI Table 10 for full list of odds ratios, SI Fig 2 and SI Table 8). We found suggestive associations with liver disease (OR=1.92, 95%CI: 1.07-3.46, p=0.03), liver fibrosis/cirrhosis (OR=3.96, 95%CI: 1.43-10.95, p=0.008), liver disease (OR=1.84, 95%CI: 1.04- 3.26, p=0.04), liver cancer (OR=3.91, 95%CI: 1.21-12.63, p=0.02), joint replacement surgeries (OR=1.93, 95%CI: 1.11-3.36, p=0.02), and any outcome (OR=1.59, 95%CI: 1.09-2.30, p=0.01), though these were not statistically significant after correction for multiple testing (p ≤ 0.0014).

PGS for hepcidin, sTfR, SF, and TIBC showed no statistically significant associations with outcomes. See SI Table 10 and SI Fig 2.

After excluding individuals with haemochromatosis at baseline, the only significant association after correction for multiple testing was that between serum iron PGS and haemochromatosis. See SI Table 9 for full list of associations in male C282Y homozygotes without a diagnosis of baseline haemochromatosis. Results remained consistent after excluding participants with relatedness to male C282Y homozygous participants (SI Table 11).

### 3.3 PGS and Disease Outcomes in Female *HFE* C282Y Homozygotes

Females with higher TSAT PGS had significantly increased odds of haemochromatosis diagnosis (OR per 1-SD increase = 1.33, 95%CI: 1.18-1.51, p=4.09×10^-6^) compared to those without a diagnosis of haemochromatosis. Similarly, higher serum iron PGS was associated with increased odds of haemochromatosis (OR=1.35, 95%CI: 1.20-1.53, p=1.60×10^-6^). Hepcidin, sTfR, serum ferritin and TIBC polygenic scores were not associated with outcomes after controlling for multiple tests (p ≤ 4×10^-6^) (SI Table 12).

Females in the highest TSAT PGS quintile were more likely to be diagnosed with haemochromatosis (OR=2.21, 95%CI: 1.50-3.25, p=0.00006), compared to those with the lowest TSAT PGS quintile. The cumulative incidence by age 80 was 45.3% [95%CI: 36.9%-52.7%] versus 23.3% [17.3%-28.8%] in the lowest quintile (p=1×10^-5^) (Fig 3A, B and SI Table 13 and 14). In Fig 3A, red points indicate associations that passed the Benjamini-Hochberg FDR threshold (p ≤ 6×10^-5^), orange points represent nominal significance (p < 0.05), and grey points are not significant (p > 0.05). The composite outcome analyzed comprised liver disease, liver fibrosis/cirrhosis, and osteoarthritis. Cumulative incidence estimates for haemochromatosis were calculated from age 40 to 80 years. Equivalent plots excluding participants with prevalent haemochromatosis are shown in SI Fig 4.

**Fig 3:**
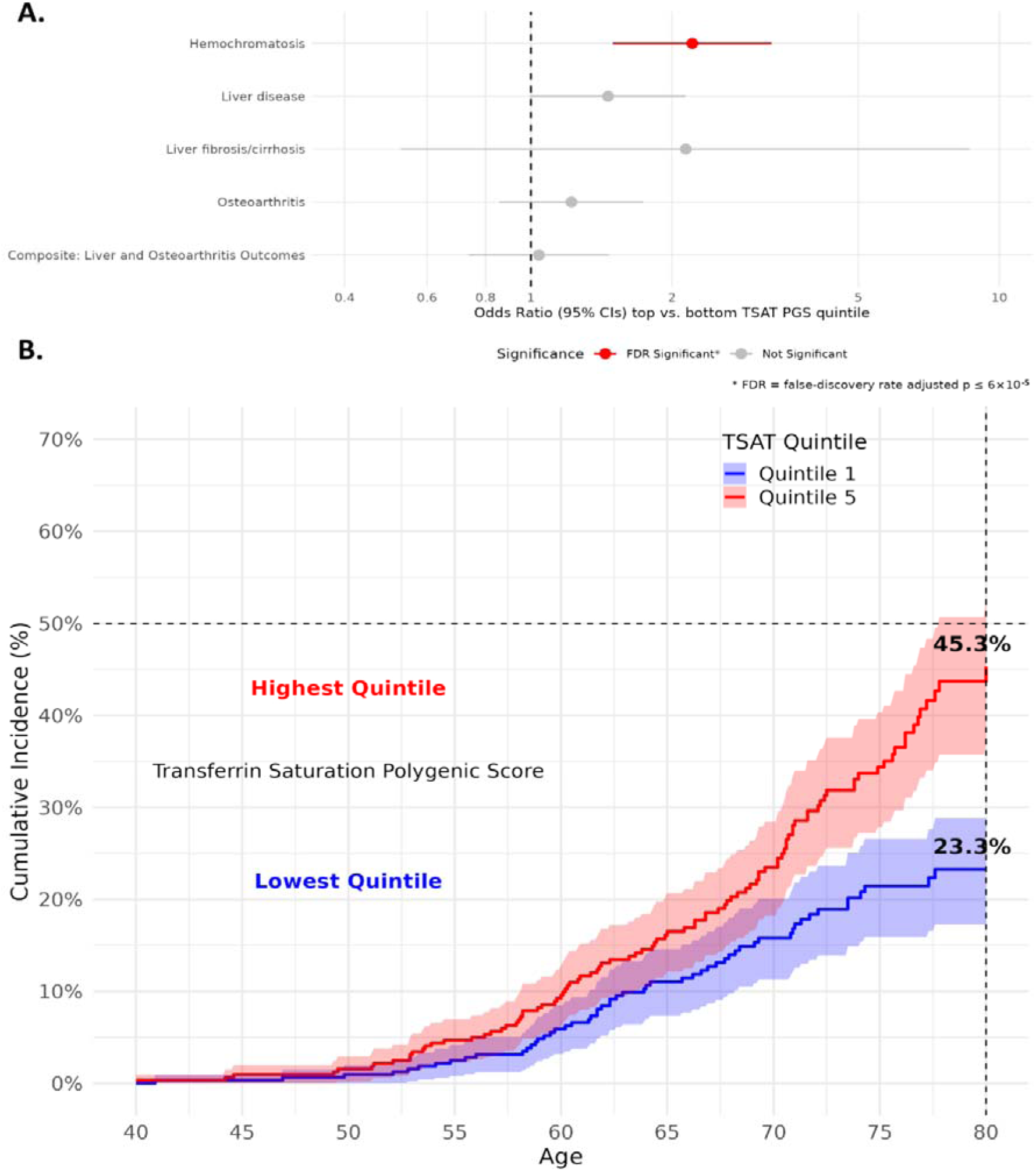
Associations between high genetically predicted transferrin saturation and clinical outcomes in female C282Y homozygotes. A) Forest plot from logistic regression analyses in male C282Y homozygotes (n=1,295) of associations between high genetically predicted TSAT (top PGS quintile) and risk of hospital-diagnosed clinical outcomes (vs bottom PGS quintile). The composite outcome includes liver disease, liver fibrosis/cirrhosis, and osteoarthritis. Red points indicate associations that passed the Benjamini- Hochberg False Discovery Rate (FDR) threshold (q < 0.05), orange points represent nominal significance (p < 0.05), and grey points are not significant. B) Cumulative incidence for haemochromatosis is plotted in the high and low TSAT PGS groups, showing the cumulative incidence of diagnoses from age 40 to 80. See SI Figure 4 for equivalent plots after excluding participants with prevalent haemochromatosis.

Similarly, females in the highest serum iron PGS quintile had increased odds of being diagnosed with haemochromatosis (OR=2.37, 95%CI: 1.57-3.57, p=3.89×10^-5^), with a cumulative incidence of 39.0% [95%CI: 31.5%-45.7%] compared to 20.5% [13.9%-26.5%] in the lowest quintile (p=0.0001) (SI Table 13 and 14 and SI Fig 3).

Hepcidin, sTfR, SF, and TIBC PGS showed no statistically significant associations with any outcomes (SI Table 13 and SI Fig 3).

In female *HFE* C282Y homozygotes, the results were largely unchanged when excluding participants diagnosed with haemochromatosis prior to baseline assessment. Haemochromatosis remained the only outcome associated with the highest quintile of TSAT and serum iron PGS (see SI Table 15 for full results and SI Fig 4 for TSAT associations without baseline haemochromatosis diagnosis). Results remained consistent after excluding female participants who were genetically related to other female C282Y homozygotes (SI Table 16).

### 3.4 Effect of serum iron and TSAT PGS on haemochromatosis incidence in non-C282Y homozygotes

In logistic regression analysis of outcome “ever diagnosed with haemochromatosis,” higher TSAT and serum iron PGS in H63D homozygous males significantly increased odds (TSAT OR per 1-SD = 2.11, 95%CI: 1.55-2.91, p=6.00×10⁻⁵; iron OR = 1.63, 95%CI: 1.21-2.23, p=0.015). Associations in other *HFE* genotype groups (C282Y heterozygotes, etc.) were not statistically significant (FDR p >0.05). In analyzes comparing highest versus lowest quintiles of PGS, no associations were significant after multiple testing correction (FDR p > 0.05).

We found similar estimates of cumulative incidence of haemochromatosis to age 80 between the top and bottom quintile of iron or TSAT polygenic score, in analyzes stratified by *HFE* genotype. For example, among C282Y heterozygous males, cumulative incidence estimates were 0.6% (95%CI: 0.3- 0.9) vs. 1.1% (95%CI: 0.7-1.5) for serum iron PGS (difference p=0.178), and 0.6% (95%CI: 0.3-0.8) vs. 0.8% (95%CI: 0.4-1.1) for TSAT PGS (difference p=0.279). These differences were not statistically significant. Similar trends of non-significant differences were seen across other *HFE* genotype groups (SI Table 17), and in analysis of female participants (difference p>0.05 for all comparisons).

### 3.5 Predictive Models for Disease Outcomes

In time-to-event (Cox) models from the baseline assessment we found that age, lifestyle factors, and genetics were statistically significant predictiors for haemochromatosis and clinical outcomes among C282Y homozygotes (Table 2). Three nested models were compared: Model 1 (age only), Model 2 (age plus lifestyle factors including alcohol intake, red or processed meat consumption, smoking status, waist-to-hip ratio [WHR], and body mass index [BMI]), and Model 3 (age, lifestyle factors, and polygenic scores [PGS]). Among male C282Y homozygotes, model C-statistic increased from 0.51 (age only) to 0.58 with lifestyle factors (p=0.003), and further to 0.60 with the addition of iron-related PGS, although the addition of PGS did not significantly improve discrimination (p=0.14). In females, the impact of lifestyle was even more pronounced, with the C-statistic increasing from 0.51 (age only) to 0.64 (age + lifestyle; p=1.05×10^-7^), and further to 0.68 when incorporating PGS (p=0.02). For liver disease in females, the model C-statistic increased from 0.56 (age only) to 0.70 (age + lifestyle; p=0.002), but remained at 0.70 with PGS (p=0.74). A similar pattern was seen for any haemochromatosis-related clinical outcome, with female C-statistics improving from 0.61 (age) to 0.66 (lifestyle; p=7.28×10⁻⁵) and to 0.66 with PGS (p=0.43). See Table 2 and SI Table 18 for full C-statistics and model comparisons across outcomes.

### 3.6 Rare Variants in *HFE* Increased Risk of Haemochromatosis

In the analysis of 28,481 variants within 2Mbp of the *HFE* gene with minor allele count ≥10 we identified 8,829 statistically significant (p < 5×10^-8^) associations with haemochromatosis diagnosis (see SI Table 19 for single variant results and SI Fig 5 for Locus Zoom plot). Of these, 4,255 had low correlation (Linkage Disequilibrium [LD]) with C282Y genotype (R^2^ < 0.01): however, none remained significantly associated with haemochromatosis diagnosis after conditioning on C282Y genotype (nominal p-values > 0.001), demonstrating the strength of association in the region due to cryptic LD with the known genotype.

In the aggregate variant analysis, we combined very rare variants (minor allele frequency <0.1%) with similar predicted functions together in “masks” such as “HFE predicted loss-of-function” (pLoF). One mask was associated with haemochromatosis after conditioning on C282Y genotype: carriers of any of 31 rare *HFE* pLoF variants had increased likelihood of haemochromatosis diagnosis (OR=14.8, 95%CIs 4.65 to 47.10, p=0.003) (see SI Table 20 for aggregate results). Other masks, such as for rare missense variants, were not significantly associated with haemochromatosis (p>0.05). Among EUR-like genetic ancestry participants, 142 carried any of the pLoF variants in the haemochromatosis-associated mask. Three individuals with a pLoF variant in *HFE* have a diagnosis of haemochromatosis in the available data (two were heterozygous for *HFE* C282Y). One of these three pLoF variants has previously been reported in ClinVar as pathogenic from a single source (chr6:26091040:AG:A is a single nucleotide deletion resulting in a frameshift: NM_000410.4(HFE):c.279del (p.Trp94fs)). We found no associations between these rare variants and risk of liver disease, liver cancer, osteoarthritis, or dementia, although analyses for these outcomes were limited by very small case numbers among variant carriers (SI Table 21). Thirty of the 31 pLoF variants in the mask were in linkage disequilibrium with *HFE* C282Y (D’ = 1), though one was not (chr6:26090975:C:T D’ = 0.07). For chr6:26091040:AG:A (the previously reported pathogenic pLoF variant) the estimated haplotype frequencies are consistent with the minor allele being out of phase with C282Y, i.e., not co-inherited on the same haplotype.

## 4 Discussion

Given the high prevalence of *HFE* C282Y in many populations and variation in clinical outcomes, it is important to identify those who may benefit from earlier or more targeted treatment. We find that male *HFE* C282Y homozygotes genetically predisposed to elevated transferrin saturation had a substantially increased risk of haemochromatosis and severe clinical outcomes, such as liver and musculoskeletal complications, compared to those with genetically predicted low transferrin saturation.

These findings suggest that genetic factors beyond *HFE* variants influence penetrance, as seen in other conditions where polygenic scores stratify patients by severity.(31,32) Incorporating genetic, demographic, and lifestyle factors into predictive models significantly improves risk prediction accuracy, therefore highlighting the complex and multifactorial nature of haemochromatosis clinical penetrance. Clinically, integrating polygenic risk assessments alongside routine *HFE* genetic testing could substantially enhance precision therapies for iron overload, enabling earlier identification of high-risk individuals and informing tailored interventions aimed at mitigating disease progression.

We also find evidence that rare predicted loss-of-function (pLoF) variants in *HFE* increased the risk of haemochromatosis in non-C282Y homozygotes. This is consistent with perturbations in this gene leading to hepcidin dysregulation, with consequent increases in iron absorption. This adds to the growing literature on other rare genetic causes of iron overload.(33,34) Three pLoF variant carriers were diagnosed with haemochromatosis, where two were also heterozygous for C282Y: the estimated haplotype frequencies were consistent with trans compound heterozygosity (the alleles being inherited separately). Although the ClinVar database contains numerous pLoF variants in *HFE* that are asserted as pathogenic for haemochromatosis, only one of the three pLoF identified here has previously been reported in ClinVar (chr6:26091040:AG:A). This highlights the need for further studies of rare causes of iron overload, which contribute to cases observed in non-C282Y homozygotes.

The present study extends previous work by incorporating longer health records follow-up (up to 30 October 2022, extending the 2021 cutoff used previously), polygenic scores for hepcidin and serum transferrin receptor (sTfR), additional clinical outcomes (associated with C282Y homozygosity in our recent study(8)), cumulative incidence estimates for clinical diagnoses up to age 80 stratified by iron polygenic score, combined clinical prediction modeling, and analysis of rare variants in HFE. Higher genetically predicted TSAT was the most consistent predictor of adverse clinical outcomes in C282Y homozygotes. An elevated TSAT is widely recognised as the most sensitive and early indicator of haemochromatosis - although the test itself is subject to biological variability and therefore lacks specificity.(35) There have been previous reports suggesting that a persistently elevated transferrin saturation might be associated with worse outcomes.(36) Yet there is broad acknowledgment that venesection/phlebotomy treatment is relatively less effective at addressing this aspect of the disease than it is in reducing stored iron (as reflected by serum ferritin), highlighting the pressing clinical need to understand the causal mechanisms in haemochromatosis.

In our previous analysis using continuous serum iron PGS, we observed modest but statistically significant associations with haemochromatosis across several non-C282Y male *HFE* genotypes, including H63D homozygotes and C282Y heterozygotes.(11) However, in the current study, we found no significant excess haemochromatosis cumulative incidence up to age 80 in participants with high TSAT or serum iron PGS within any *HFE* genotype group except C282Y homozygotes. These findings suggest that at the population level, common non-HFE serum iron-associated genetic variants only meaningfully impact haemochromatosis incidence in *HFE* C282Y homozygotes. This is especially evident in males, where haemochromatosis cumulative incidence reached 55.4% in the highest TSAT PGS quintile compared to 38.0% in the lowest.

While both serum iron and TSAT PGS were significantly associated with a haemochromatosis diagnosis in both male and female *HFE* C282Y homozygotes in our 2022 paper, their associations with broader morbidity, including joint replacement surgeries, dementia, and non-Alzheimer’s dementia, were not previously analysed. In the current study, both TSAT and serum iron PGS were associated with increased likelihood of liver complications, with stronger and more consistent effects observed for TSAT within male C282Y homozygotes. We found that TSAT and serum iron PGSs are highly correlated due to partially overlapping genetic variants. Clinically, TSAT is preferred over serum iron as it better reflects available circulating iron and is less affected by short-term variation,(35,37) supporting its prioritisation as the primary polygenic modifier in risk models. The stronger effect estimates of the TSAT PGS highlight its value in predicting liver and musculoskeletal outcomes in male C282Y homozygotes.

Another notable finding was the absence of strong associations with dementia outcomes. Although previous UK Biobank studies have reported increased risk of dementia associated with excess iron,(22) also within male C282Y homozygotes,(7) neither TSAT nor serum iron PGSs were predictive of these outcomes. This may reflect differing biological mechanisms or limited case numbers, suggesting iron-related polygenic effects primarily influence earlier-stage rather than end-stage complications.(7,8)

Despite the robust associations between TSAT and serum iron PGSs and clinical outcomes, several polygenic scores including those for SF, TIBC, sTfR, and hepcidin showed minimal or no associations. These findings are consistent with those from our previous study.(11) The weaker performance of these PGS biomarkers may reflect their susceptibility to non-genetic influences. Serum ferritin is an acute-phase reactant,(38) and TIBC levels may inversely reflect iron burden but lack direct causality in iron toxicity.(39)

The hepcidin PGS was also not robustly associated with clinical outcomes. While hepcidin is a key regulator of systemic iron homeostasis(40) and a target of emerging therapeutics (e.g., Rusfertide(41)), the polygenic score used in this analysis was derived from a GWAS of serum hepcidin levels, which may not fully capture the regulatory activity of hepcidin within specific tissues. The hepcidin PGS was also not robustly associated with outcomes. While hepcidin is a key regulator of systemic iron homeostasis,(40) the PGS used here may not fully capture regulatory activity within specific tissues, and limited GWAS sample sizes may have reduced power.(26) In the context of *HFE* haemochromatosis, where *HFE* and TMPRSS6 are major drivers of downstream hepcidin suppression, genetic variation in hepcidin itself may play a smaller role in modulating disease risk.

Predicting disease outcomes in haemochromatosis remains an important challenge. Combining genotypic risk factors with lifestyle factors and possibly the development of more sensitive biomarkers of disease and offers the most comprehensive approach to identifying individuals at greatest lifetime risk - and who would benefit most from targeted treatment.

### 4.1 Strengths and Limitations

A major strength of this study is the large sample of *HFE* C282Y homozygotes, enabling robust sex-stratified estimates and cumulative incidence modeling of clinical outcomes. Extended follow-up of hospital inpatient diagnoses allowed assessment of penetrance to clinical outcomes up to age 80. Compared to our previous publication, this study includes more outcome events and additional polygenic scores, offering broader insight into genetic modifiers of clinical risk in haemochromatosis and related outcomes.

This is also the first study, to evaluate polygenic scores for hepcidin and sTfR in relation to clinical outcomes specifically among C282Y homozygotes. While associations for these markers were generally weak, their inclusion provides a broader view of the potential regulatory pathways influencing penetrance. However, limitations remain. The hepcidin score was based on circulating serum levels rather than tissue-specific activity, which may limit interpretation. While predictive models improved when polygenic and lifestyle factors were included, overall discrimination remained moderate, and findings will require replication in other community-based cohorts. Lastly, the GWAS studies that identified the variants used to derive the iron-status polygenic scores were conducted in general European ancestry populations and did not specifically include *HFE* C282Y homozygotes.(26,27) Thus, we assume that the genetic effects observed in these broader populations are consistent within C282Y homozygotes.

The generalisability of findings may be limited by the UK Biobank healthier volunteer bias and incomplete primary care data coverage, which may lead to underestimation of clinical penetrance. Although associations with TSAT and serum iron polygenic scores were consistent and biologically plausible, predictive performance remained moderate, and external validation will be needed. Additionally, we did not have direct measures of TSAT in UK Biobank data, so we could not evaluate whether the TSAT polygenic score provides additional clinical value beyond actual blood measurements. Lastly, the hepcidin polygenic score, based on serum levels, may not fully capture its regulatory role in iron metabolism.

## 5 Conclusion

To conclude, genetic and lifestyle factors significantly influence clinical penetrance in C282Y homozygous individuals. Polygenic scores for transferrin saturation and serum iron predict haemochromatosis and related outcomes. Rare, large-effect variants in *HFE* likely further modify risk, and may reflect novel causes of iron overload in non-C282Y homozygotes. Lifestyle factors, such as alcohol intake, diet, smoking, and obesity, further increase outcome predictability. Our findings highlight the need for personalised screening strategies and targeted management of HFE-related iron overload in C282Y homozygotes.

## Data Availability

Data are available on application to the UK Biobank (www.ukbiobank.ac.uk/register-apply).

## Acknowledgements

This research was conducted using the UK Biobank resource, under applications 14631 and 103356. We thank the UK Biobank participants and coordinators for the dataset. This work used data provided by patients and collected by the NHS as part of their care and support. Copyright © (2025), NHS England. Re-used with the permission of the NHS England and UK Biobank. All rights reserved. This research also used data assets made available by National Safe Haven as part of the Data and Connectivity National Core Study, led by Health Data Research UK in partnership with the Office for National Statistics and funded by UK Research and Innovation. This study was supported by the National Institute for Health and Care Research Exeter Biomedical Research Center. The views expressed are those of the authors and not necessarily those of the NIHR or the Department of Health and Social Care. For the purpose of open access, the author has applied a ‘Creative Commons Attribution (CC BY)’ licence to any Author Accepted Manuscript version arising from this submission.

Funding: JA has an NIHR Advanced Fellowship (NIHR301844). The University of Exeter supports ML, LP. GH is supported by the Medical Research Council [grant number UKRI327].

## 6 Conflict of interest

None declared.

## 7 Data sharing

Data are available on application to the UK Biobank (www.ukbiobank.ac.uk/register-apply).

## 8 Author Contributions

ML: Conceptualization, Data Curation, Formal Analysis, Investigation, Visualization, Writing - Original Draft. JD: Methodology, Writing - Original Draft. RB: Data Curation, Formal Analysis, Investigation, Writing - Review & Editing. GH: Data Curation, Software, Writing - Review & Editing. AW: Data Curation, Software, Writing - Review & Editing. CW: Supervision, Writing - Review & Editing. JS: Conceptualization, Writing - Original Draft Preparation. JA: Conceptualization, Funding Acquisition, Investigation, Methodology, Supervision, Writing - Original Draft Preparation, Writing - Review & Editing. LP: Conceptualization, Funding Acquisition, Data Curation, Formal Analysis, Investigation, Methodology, Project Administration, Supervision, Writing - Original Draft Preparation, Writing - Review & Editing.

## Notes

### Competing Interest Statement

The authors have declared no competing interest.

### Author Declarations

The Ethics Committee of the North West Multicentre Research Ethnics Committee (UK) gave ethical approval for the the UK Biobank study (reference number: 11/NW/0382). Access to the data for this work was granted under UK Biobank approved applications 14631 and 103356.

