## Supplementary Information for "Genetic and lifestyle modifiers of haemochromatosis-related clinical outcomes in HFE C282Y homozygotes: prospective cohort study in UK Biobank"

Lucas et al. 2025

**Supplementary Information**

### Supplementary Methods

#### Polygenic Score for Iron Biomarkers

Supplementary Table 2 includes the genetic variants used to create the polygenic scores for each trait. The position is in human genome build 38. We used R package {ukbrapR} v0.3.5 to create the polygenic scores in the UK Biobank Research Analysis Platform (<https://github.com/lcpilling/ukbrapR>). This uses `bgenix` (<https://enkre.net/cgi-bin/code/bgen/doc/trunk/doc/wiki/bgenix.md>) to extract the variant calls from the UK Biobank whole genome sequencing DRAGEN variant calls (BGEN format, field:24309). Then `plink2` (<https://www.cog-genomics.org/plink/2.0/>) converts the subsetted BGEN file to BED format. Then `plink` v1.9 allele scoring function (<https://www.cog-genomics.org/plink/1.9/score>) is used to create the weighted allele score.

### SI Tables

For supplementary tables, see accompanying Excel file.

### SI Figure 1

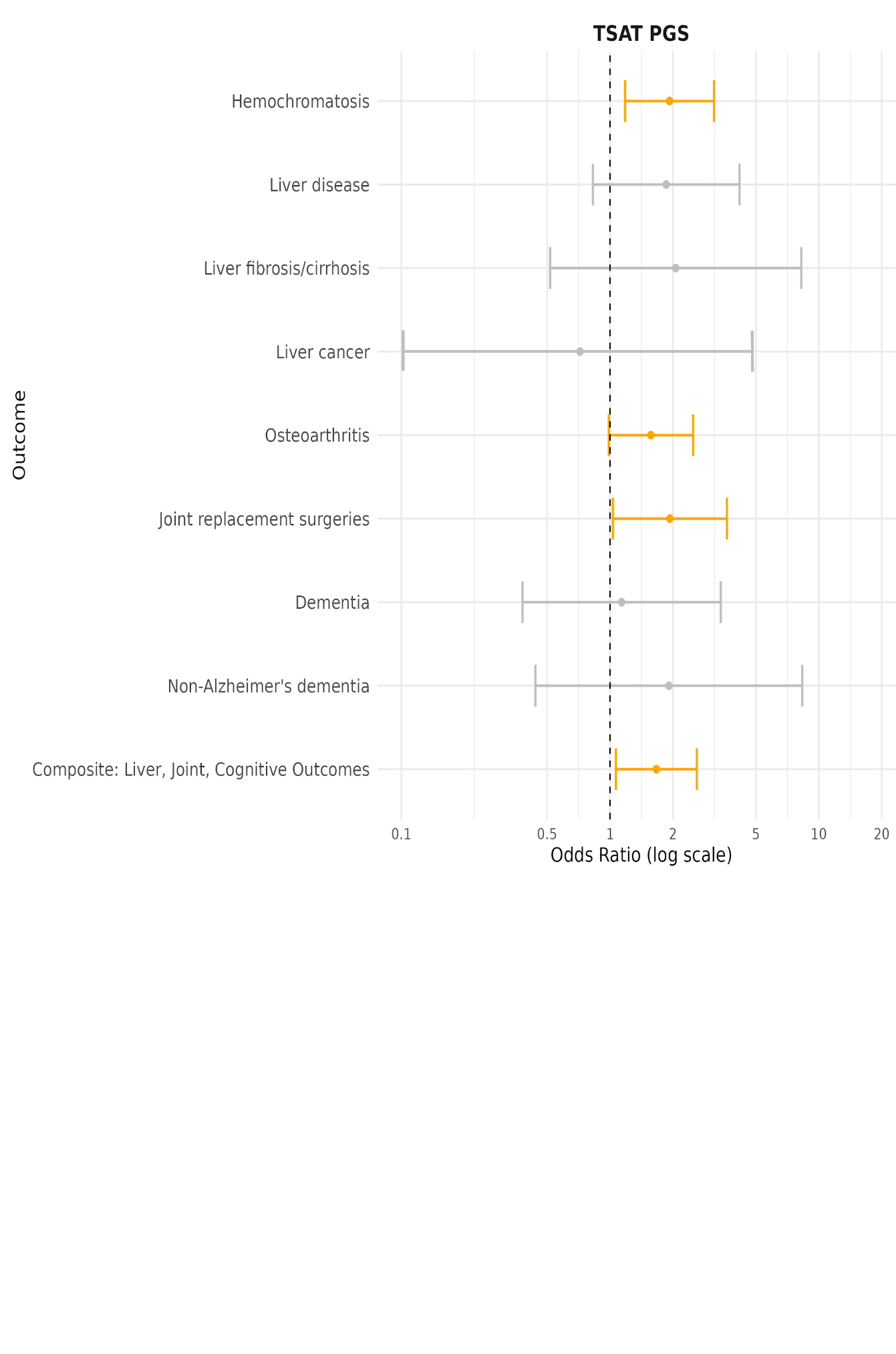

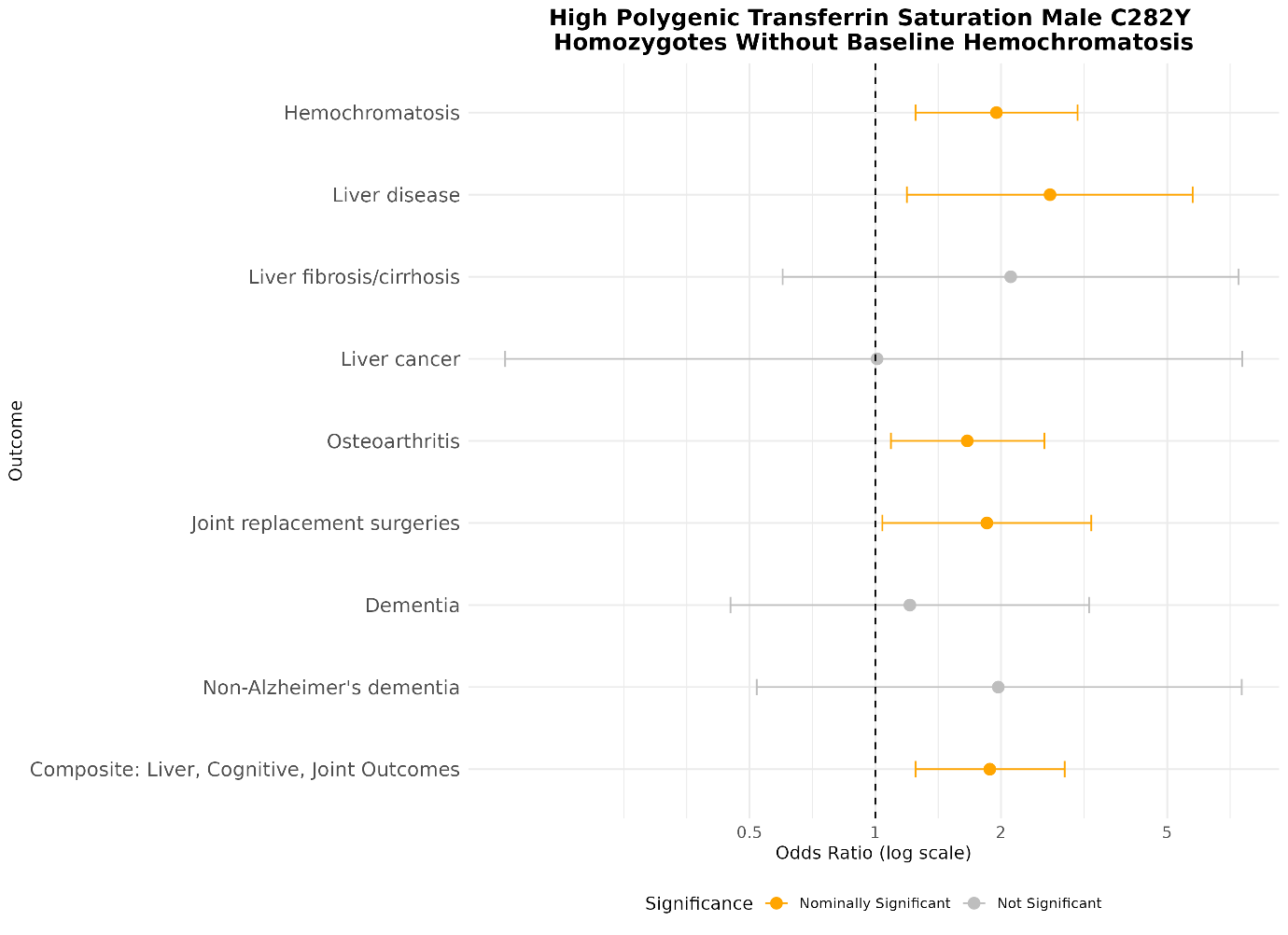

Associations between the highest quintile of TSAT polygenic biomarker and outcomes in C282Y homozygous male UK Biobank participants, without baseline hemochromatosis

### SI Figure 2

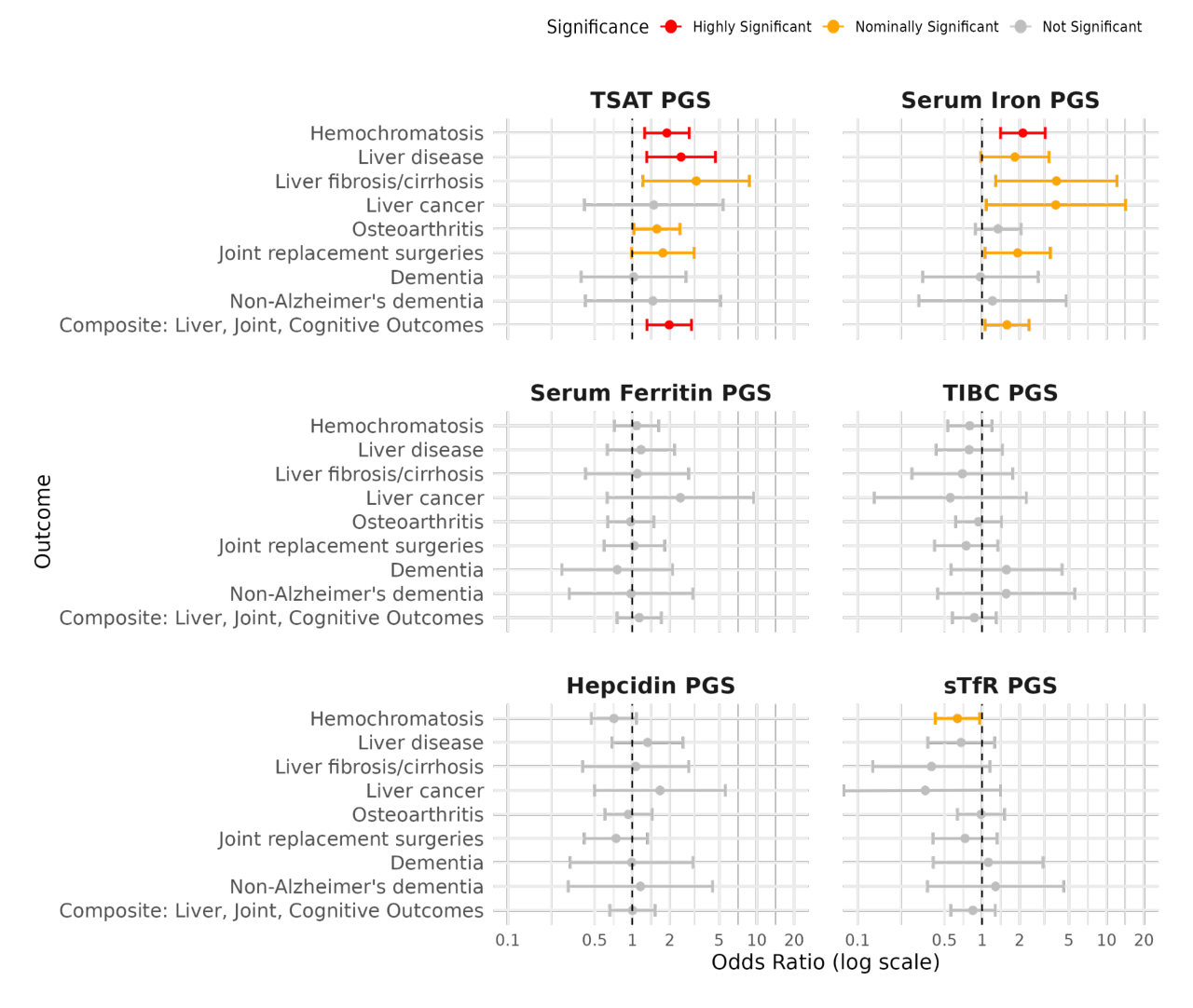

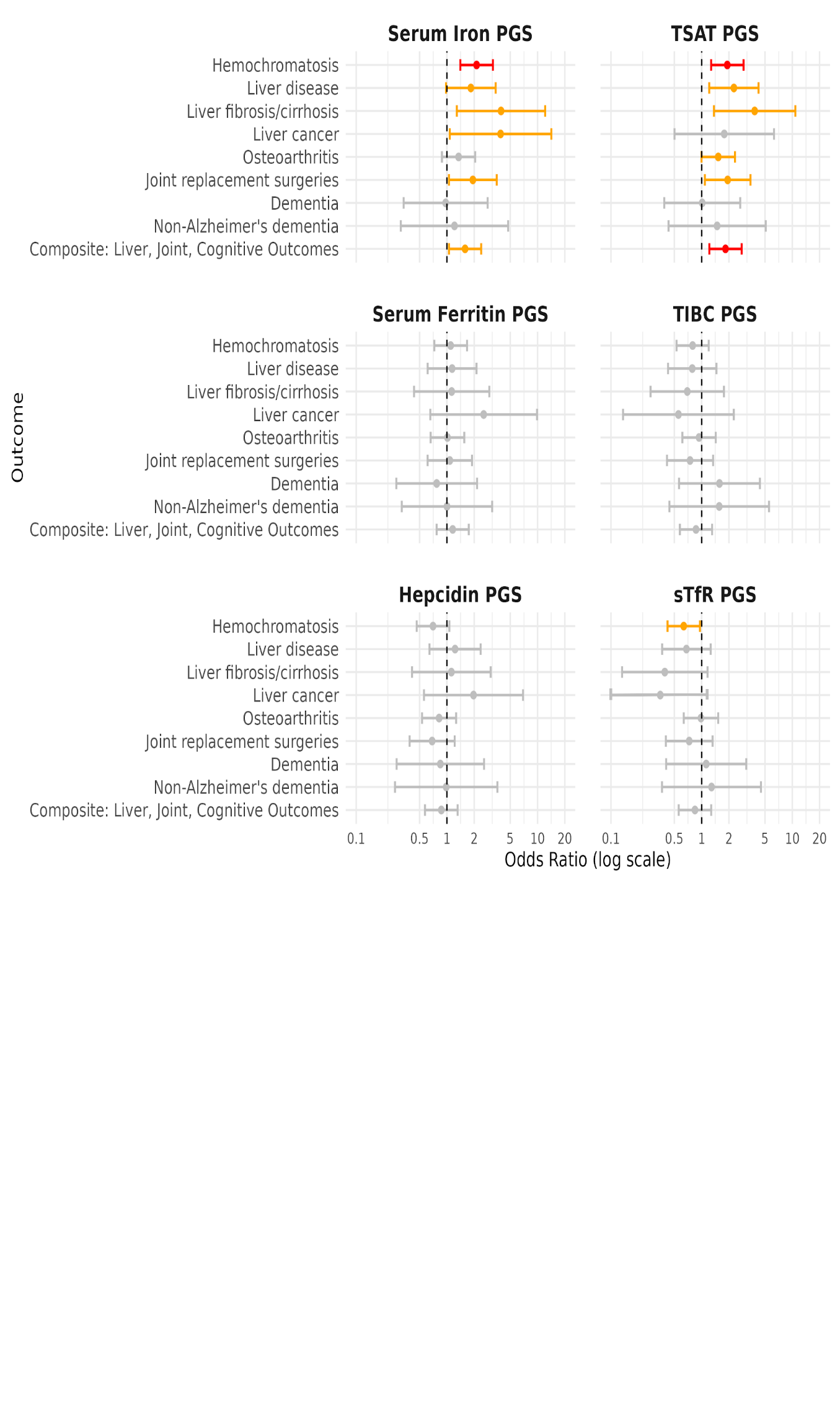

Associations between the highest quintile of iron polygenic biomarkers and outcomes in C282Y homozygous male UK Biobank participants.

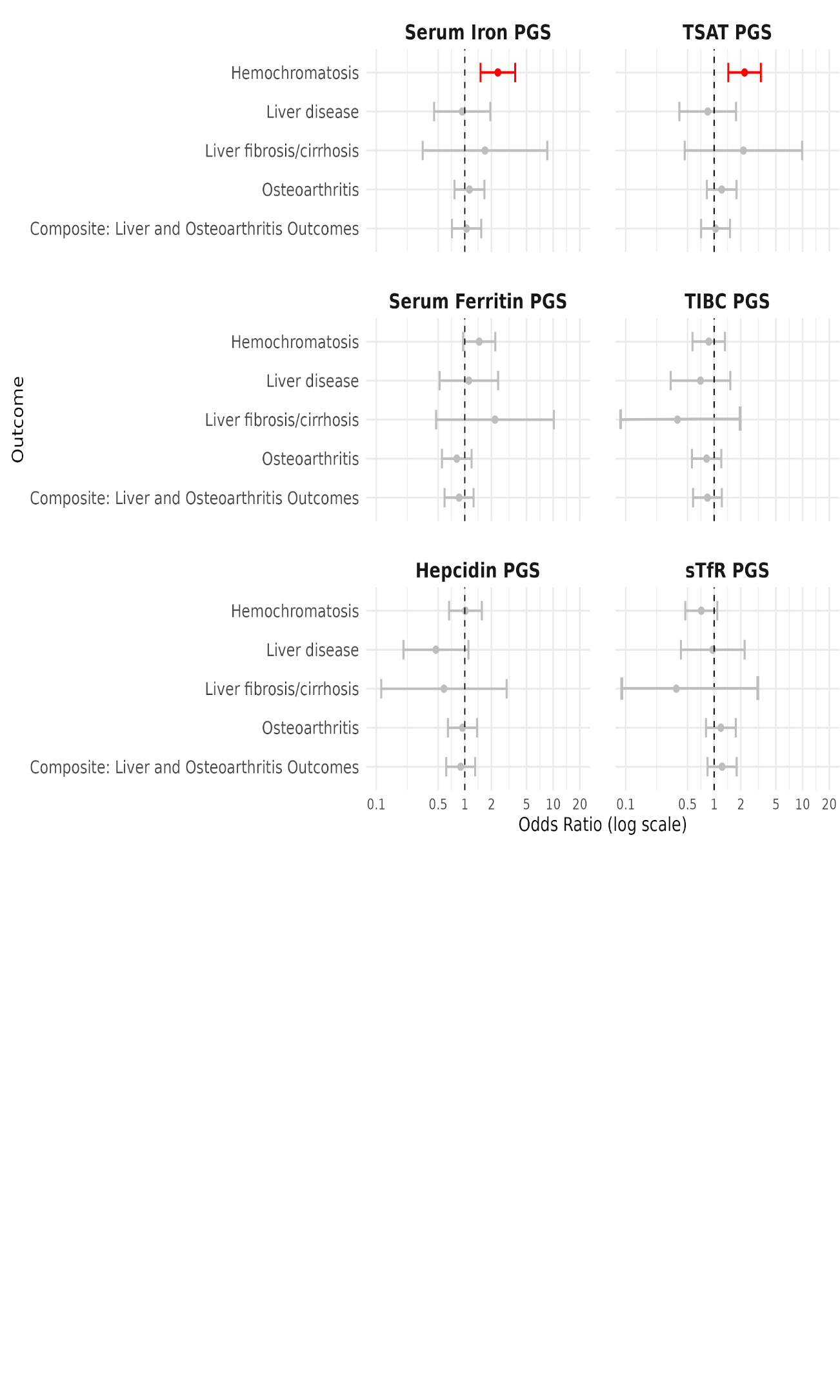

### SI Figure 3

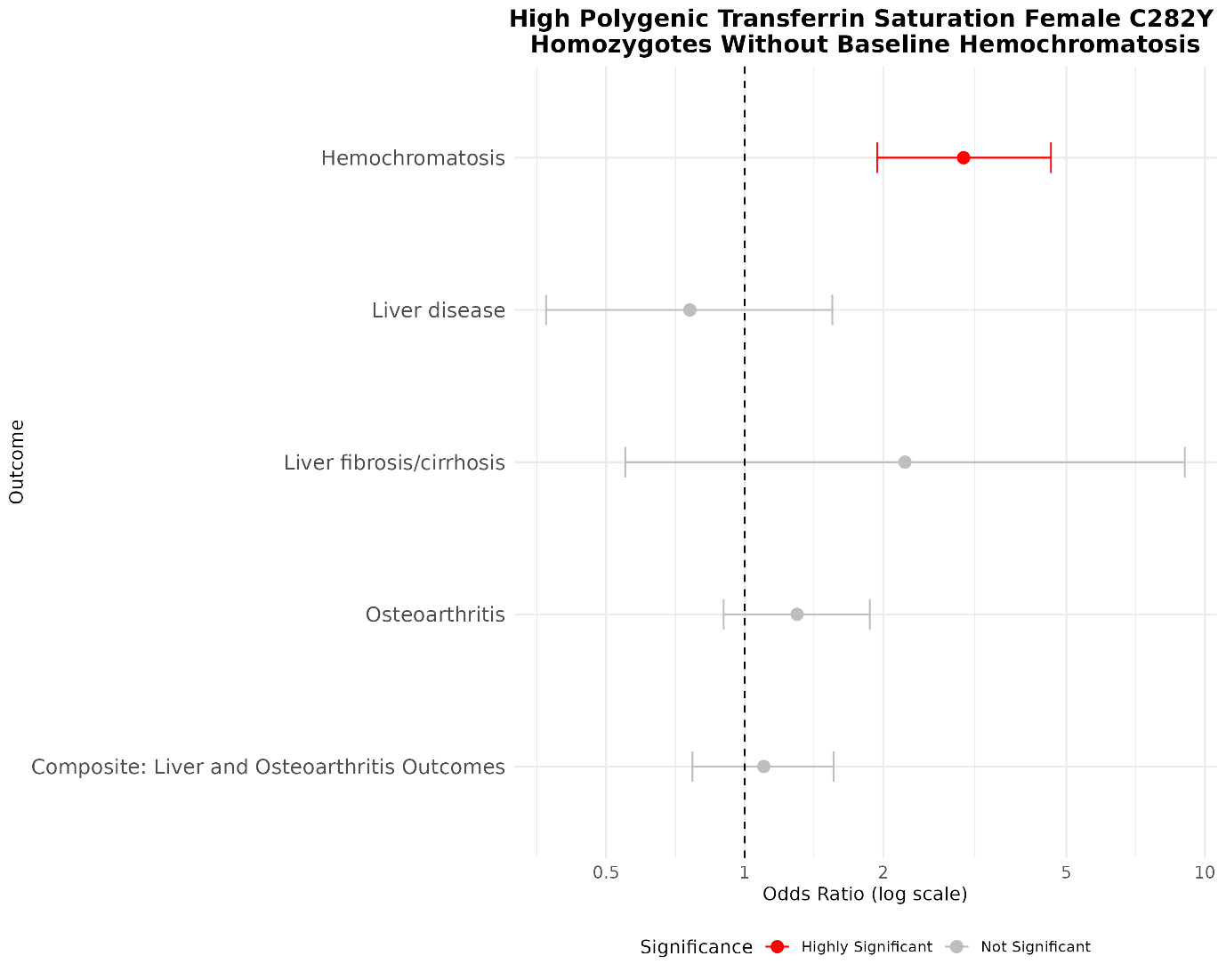

Associations between the highest quintile of iron polygenic biomarkers and outcomes in C282Y homozygous female UK Biobank participants

### SI Figure 4

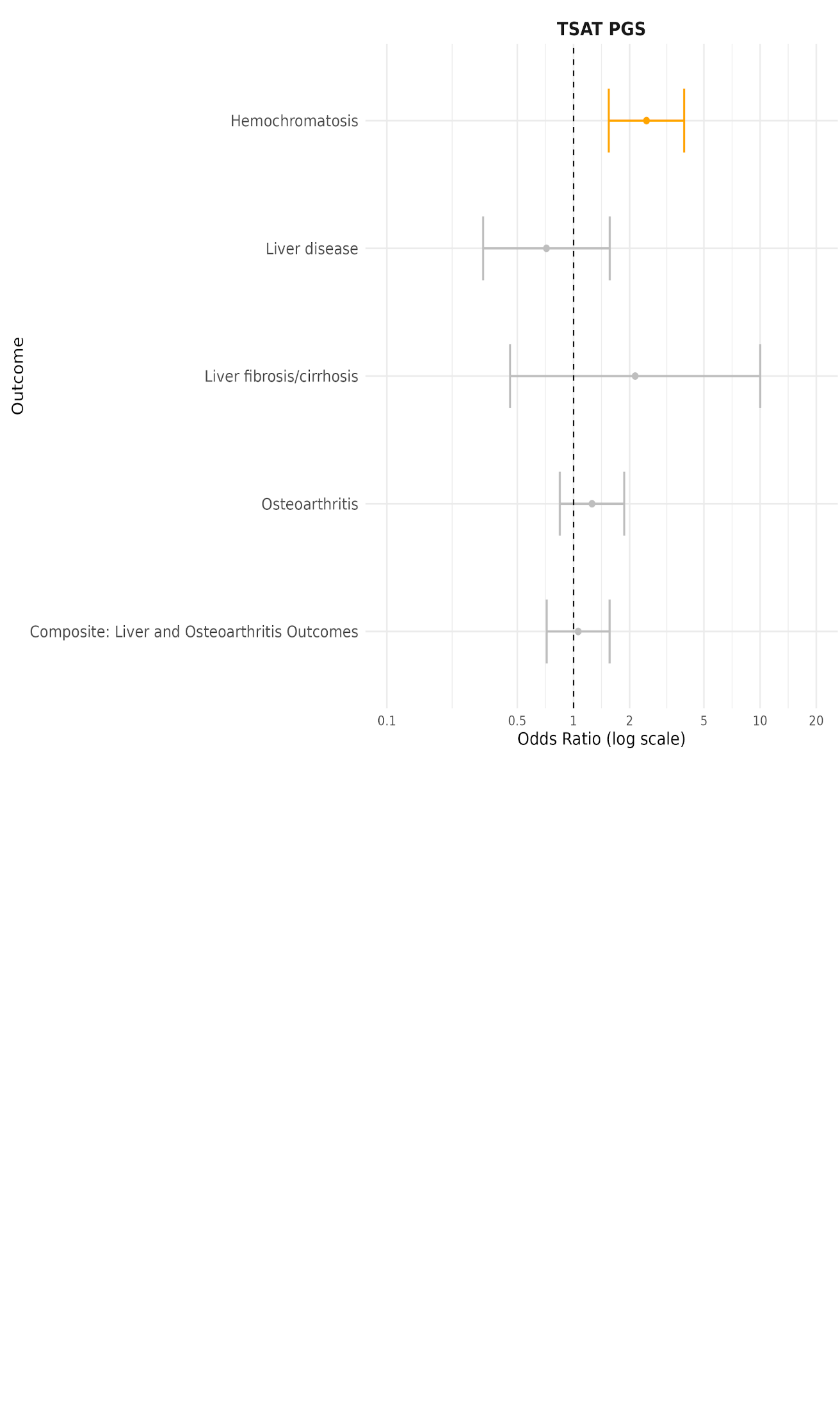

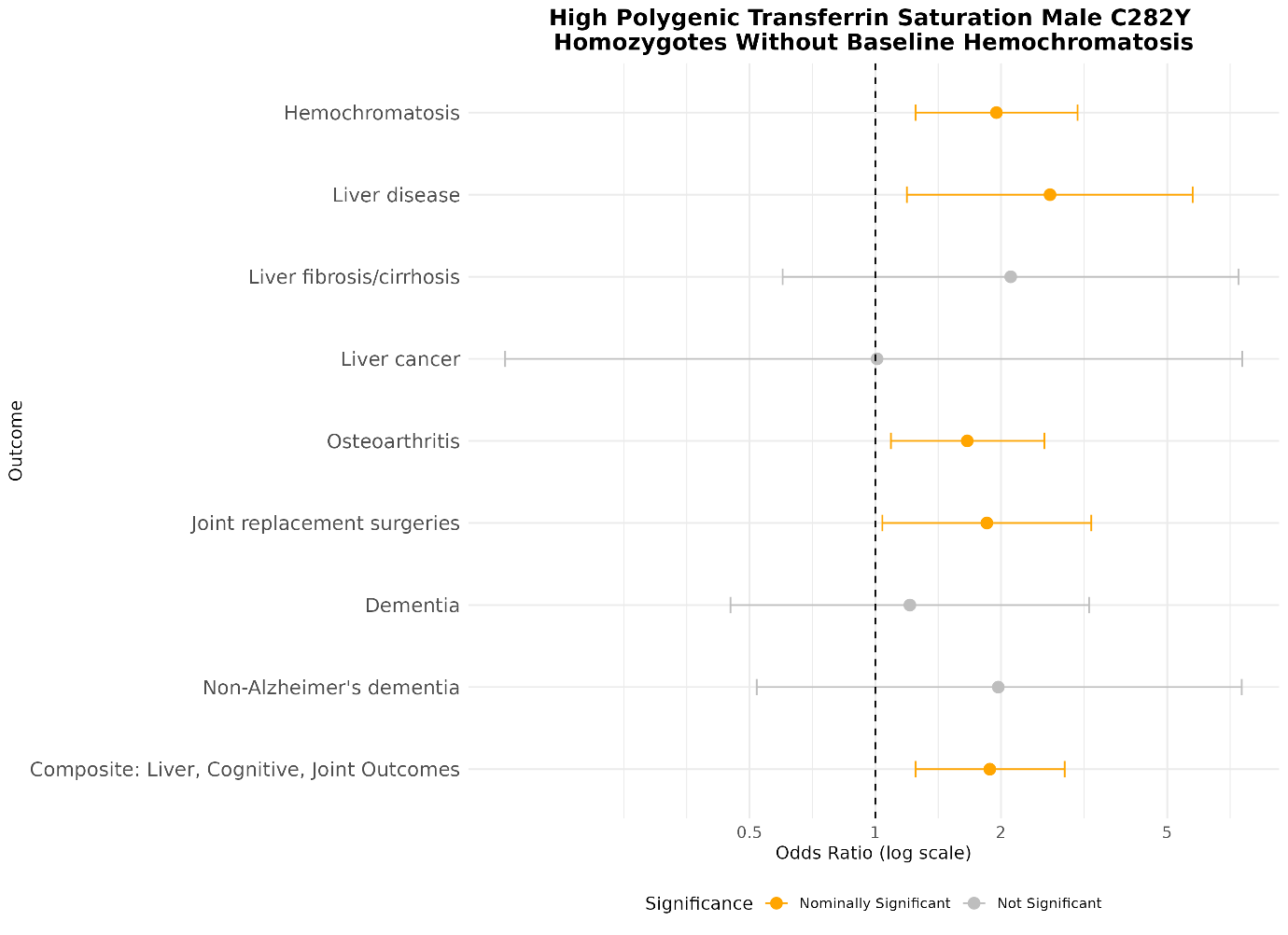

Associations between the highest quintile of TSAT polygenic biomarker and outcomes in C282Y homozygous female UK Biobank participants, without baseline hemochromatosis.

### SI Figure 5

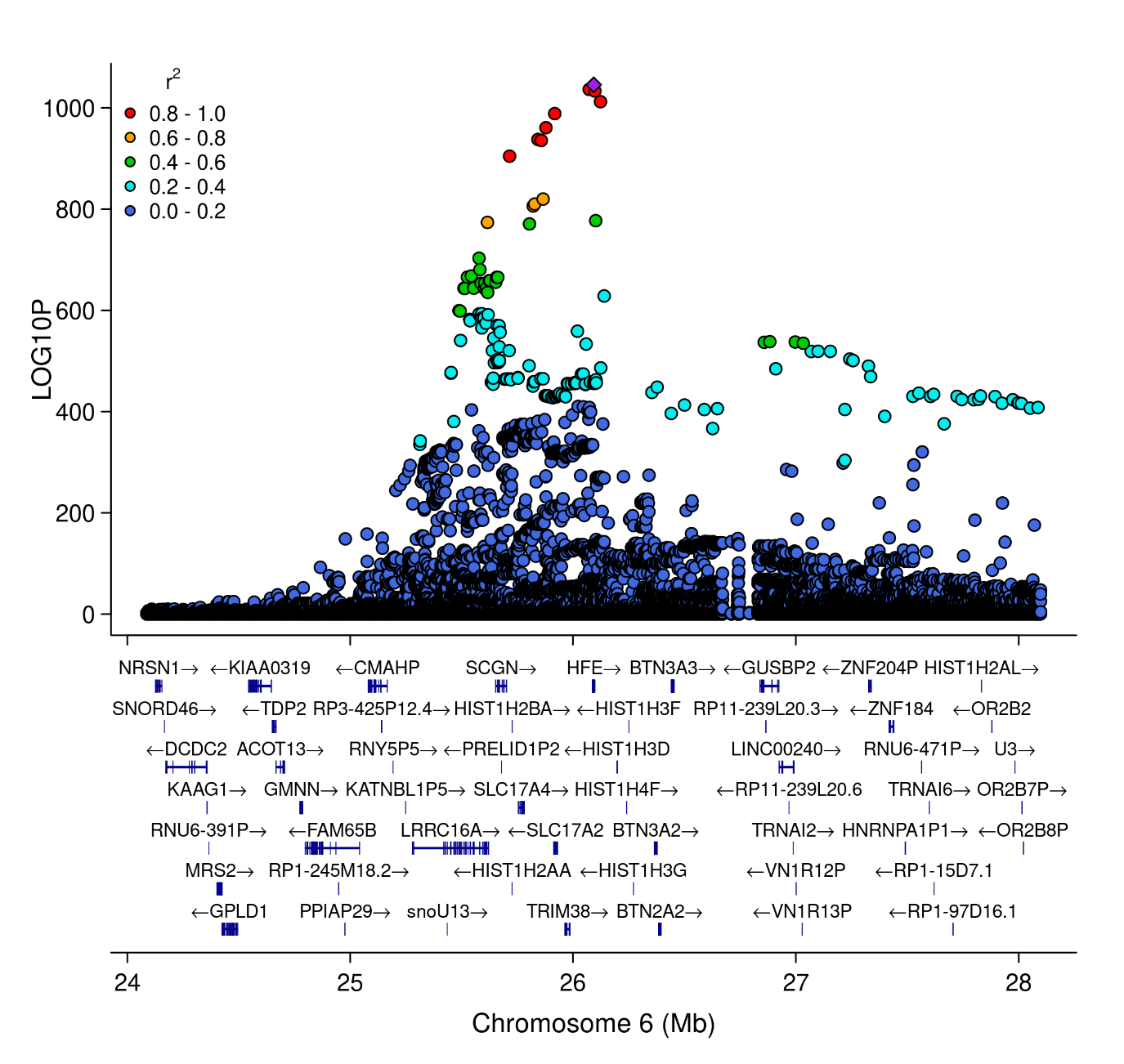

LocusZoom plot of *HFE* region on chromosome 6. Each point is a genetic variant: the x-axis shows it’s location on chromosome 6, and the y-axis is the strength of association with haemochromatosis diagnosis (-log_10_ *p*-value). The points are coloured by their correlation coefficient (R^2^) with *HFE* C282Y (rs1800562), coloured purple, in the UK Biobank EUR-like participants. Gene names and positions are indicated below the points, for reference. R package {locuszoomr} v0.3.8 was used to generate the plot (<https://github.com/myles-lewis/locuszoomr>).
